# Tobacco industry interference and WHO FCTC Article 5.3 implementation in Aotearoa New Zealand: A qualitative analysis

**DOI:** 10.64898/2026.02.24.26346935

**Authors:** Melissa-Jade Gregan, Janine Wiles, Vili Nosa, Erena Wikaire, Peter J. Adams

## Abstract

**Background:** Article 5.3 of the WHO Framework Convention on Tobacco Control requires Parties to protect policies from tobacco industry interference, yet implementation remains weak internationally. Aotearoa New Zealand’s (Aotearoa NZ) is seen as a leader in tobacco control, yet little is known about its implementation of Article 5.3 protections. This study examines these protections as well as existing transparency measures in light of the 2024 repeal of world-leading tobacco control policies.

**Methods:** Interviews with current and former: public health experts, politicians, officials and political journalists, and analysis of key texts.

**Results:** Aotearoa NZ’s Article 5.3 implementation and scope is constrained, leaving invisible and exploitable paths of influence. Public health experts argued protections have been neglected from the start. Politicians were unaware of Article 5.3 obligations, and reported limited guidance on industry interactions. These gaps are compounded by non-existent lobbying laws and ill-equipped transparency measures.

**Conclusion:** Despite the country’s reputation for strong tobacco controls, structural policy and implementation failures leave Aotearoa NZ’s health policies vulnerable to industry interference. Aotearoa NZ and other Parties should consider institutionally embedding comprehensive Article 5.3 protections to safeguard policy decisions from tobacco industry influence.

**WHAT THIS PAPER ADDS:** *What is already known on this topic:* Tobacco industry interference remains the biggest barrier to tobacco control policies, with evidence consistently identifying gaps in Parties implementation of Framework Convention on Tobacco Control Article 5.3 protections. Parties often rely on pre-existing measures such as lobbying laws.

*What this study adds:* This is the first study examing Aotearoa NZ’s implementation of Article 5.3. It shows that despite its reputation as a tobacco control leader, implementation is severely limited and pre-existing measures are inadequate, enabling a system in which industry interference can go on unseen.

*How this study might affect research, practice or policy:* By identifying how structural policy gaps enable industry interference, this study highlights the need for comprehensive institutional embedding of Article 5.3 protections across government, and consideration of its codification into law.

## BACKGROUND

Tobacco industry interference undermines public health policymaking globally, representing one of the most significant barriers to effective tobacco control [1]. Article 5.3 of the World Health Organization Framework Convention on Tobacco Control (WHO FCTC) recognises this and requires Aotearoa NZ and other Member States (‘Parties’) of the WHO FCTC to protect public health policy from the “commercial and other vested interests of the tobacco industry” [2]. Comprehensive implementation guidelines adopted in 2008 set out eight overarching recommendations (each supported by a set of specific recommended measures, 34 in total), to be implemented across all “branches of government,” [1] including the executive (Cabinet), legislature (Parliament), and judiciary (courts).

Reviews of Article 5.3 implementation reveal sizable implementation gaps globally [3–5]. For example, Fooks et al.’s [4] comprehensive assessment found 83 percent of 155 Parties had employed less than a third of the specific measures, with many Parties relying on pre-existing mechanisms such as lobbying regulations rather than tobacco-specific protections. However, unlike other jurisdictions, Aotearoa NZ does not regulate lobbying or its disclosure.

In November 2023, Aotearoa NZ’s newly elected centre-right coalition government shocked global health advocates by announcing the repeal of world-leading tobacco control laws in their coalition agreements, and enacting the repeal in March 2024 [6–8]. The repealed policies included reducing the nicotine content of tobacco to non-addictive levels, making it illegal to sell tobacco products to anyone born after 2009 (a ‘smokefree generation’), and reducing tobacco retail outlets from approximately 6,000 to 600 [9]. The government argued that the repeal was a requirement of their three-party coalition agreement and the policies would have had “a significant impact” [10] on promised tax cuts (by reducing tobacco excise income), and “increased [the] black market” trade in tobacco [11].

The repeal has profound equity implications. Tobacco-related harm is distributed inequitably with Māori (the Indigenous peoples of Aotearoa NZ) and Pacific peoples (those in Aotearoa NZ born in, or who culturally identify with, Pacific Island nations), and those living in low socioeconomic areas disproportionately impacted due to external factors including industry targeting, inequitable cessation support, and the impact of ongoing racism and colonisation [12,13]. In 2023/24, approximately 14.7 percent of Māori adults and 12.3 percent of Pacific adults smoked daily, compared to 6.9 percent of the overall adult population, and 13.9 percent of adults living in the most deprived neighbourhoods smoked daily compared to 2.5 percent of those in the least deprived neighbourhoods [12]. The WHO FCTC recognises the disproportionate burden of tobacco use among Indigenous peoples and requires Parties to develop culturally appropriate tobacco control programmes with Indigenous participation [14,15]. The repealed policies had embedded Māori health aspirations through active engagement and establishment of a Māori Governance group [16]. Government was advised that the repeal would have a “disproportionately negative impact” on these populations [17]. Modelling forecast that the policies would have achieved the Smokefree 2025 goal by 2027 for Māori, compared to 2061 without them [16].

This policy reversal occurred within a wider context of sustained tobacco-industry influence. Media reports of a leaked 2017 internal PMI document, indicated deliberate industry targeting of specific political parties to secure favourable regulation for its “smoke-free products” [18]. Internal documents from 2019 of US vaping company, JUUL, held in the Truth Tobacco Industry Documents Library, allege that one of the political parties later included in the 2023 coalition government had an established “relationship with PMI [Philip Morris International]” [19]. Two former senior party officials from this same political party took up external relations roles at PMI from 2019 onwards [18,20,21], one of whom is described by a senior Minister of the coalition political party as a “long term associate and friend” [22]. Furthermore, this Minister told reporters they knew nothing about the WHO FCTC and would not give it “one iota of attention” [23], and admitted the party took policy “soundings” from the official in their PMI role [22]. Then, in July 2024, the coalition government approved a 50 percent tax cut on PMI’s heated tobacco products, on the advice of another Minister from this political party [24]. Officials had warned government the cut would solely benefit PMI, which holds a monopoly on these products in Aotearoa NZ [25].

While Article 5.3 implementation has been examined internationally, Aotearoa NZ’s implementation remains under-explored. This article aims to address this gap. We sought to understand the nature and extent of implementation of Article 5.3 and the constraints on tobacco industry influence in policy processes. Understanding these mechanisms is essential both for strengthening Aotearoa NZ’s protections and informing best-practice implementation of Article 5.3 globally, particularly in jurisdictions without lobbying disclosure or protections.

## METHODS

We drew on institutional ethnography (IE), a critical qualitative research approach that examines how key institutional ‘texts’ shape people’s work practices [26,27]. Texts refers to replicable written or electronic instructional messages that are often administrative, managerial, or legislative in nature [26,28]. The term ‘institution’ refers to organised systems of activity structured around a particular social function: we focused on the social institution of political lobbying in Aotearoa NZ, specifically interactions between politicians and interest groups advocating either for public health or for industry priorities. IE enables researchers to trace complex social relations largely invisible to those outside the institution, making it ideal for studying lobbying practices in contexts without mandated lobbying disclosures [29]. The findings presented here are drawn from a larger study of industry lobbying in Aotearoa NZ that also included alcohol, gambling and ultra-processed food industries [30].

### Data collection

Data included key informant interviews and relevant texts. Study participants, known as informants in IE, included: current and former politically active public health experts (n=11 of 14 invited), of which five were Māori and Pacific public health experts (n=5 of 8 invited); and current and former members of Parliament (MPs), Ministers, officials, and political journalists (n=10 of 31 invited). Public health experts were identified and contacted through our professional networks. Political informants were identified based on one or more criteria: involvement in relevant policy areas (e.g. as Ministers, select committee members, or advisors); documented public commentary on relevant policy areas; or documented semi-regular interaction with relevant interest groups (evidenced by ministerial diary entries or select committee records). Ministry of Health officials did not respond to invitations to participate. To support informant confidentiality in Aotearoa NZ’s small and interconnected policy environment, we exclude all identifying characteristics. This includes referring to both MPs and ministers as ‘politicians’.

Interviews were semi structured and took place between March 2021 and May 2023, either in person or online (using Zoom) according to Aotearoa NZ’s COVID-19 restrictions and informant preference, and primarily lasted between 40 and 60 minutes (three lasted up to 120 minutes). This data collection period preceded the political changes in Aotearoa NZ’s tobacco control landscape described above. Public health experts were asked to recount attempts to access and interact with policymakers, and about any direct experience with Article 5.3. Political informants were asked about interactions with public health and industry actors and what guided these interactions. Key texts were identified through interviews or proactive searches, and included: the WHO FCTC and Article 5.3 implementation guidelines; Aotearoa NZ Ministry of Health biennial implementation reports to the WHO FCTC Secretariat; agency registers of meetings with tobacco industry actors; political correspondence and existing official information requests; parliamentary submissions; and government transparency documents (e.g., Official Information Act 1982 and ministerial diary summaries).

### Data analysis

Institutional ethnographic data collection and analysis is iterative, and aims to explicate the structural organisation of people’s experiences. This is achieved through text-talk-text approaches [31] in which researchers move back and forward between texts and informants’ accounts of actual work practices to trace how work practices are shaped by relevant institutional texts [32]. Beyond explicitly identified texts, connections are identified through “direct and indirect references to institutional practices such as meetings, appointments, schedules, policy and rulemaking” [32,33]. We used NVivo 12 software to categorise data into work practices (e.g., ‘relationship building’, ‘information provision’, ‘hospitality and gifts’), and texts (e.g., ‘ministerial diary summaries’, ‘legislation’, ‘policies’) to enable us to map connections between actions and institutional texts.

## RESULTS

Despite being considered a leader in tobacco control globally, on closer analysis Aotearoa NZ’s approach to protecting public health policies from tobacco industry interference reveals substantial holes in scope, awareness, and effectiveness.

### Neglected from its inception

Public health experts interviewed stated that Article 5.3 guidelines have been neglected since their introduction. One recalled the guidelines were: “very much a non-event in New Zealand. Even in the Ministry of Health.” A key factor considered to contribute to this neglect was Article 5.3 not being codified into Aotearoa NZ law. This expert argued that without legislative embedding, “government departments don’t really think that an international treaty is hard law,” rendering the guidelines what they described as “soft law.” The public health expert contended that the government needed to embed the WHO FCTC into law with sufficient “teeth”:

> [Then] it only takes an OIA [request under the Official Information Act 1982] or a media enquiry or in the extreme case a judicial review, to expose that the law is not being carried out. (Public health expert)

Public health informants also described little engagement with the Article 5.3 guidelines within the Ministry of Health. For example, one stated, “hardly anyone read about them [laughs] let alone thought about them.” Another expressed frustration, stating:

> What’s the point of [Article] 5.3 if you’re not actually adhering to it? It should be a hell of a lot tighter than it is. They [tobacco industry] shouldn’t even be able to get in the front door. (Public health expert)

Our analysis of Aotearoa NZ’s biennial WHO FCTC implementation reports supports this pattern. In 2010, two years after the guidelines were published, the Ministry of Health reported it was still considering “how to implement the Article 5.3 guidelines” [34]. When public health informants discussed Article 5.3 with officials at the time, they reported being told “there was transparency there already [that would] fit with 5.3.” The transparency mechanisms officials were referring to remains unclear from available records.

### Constrained scope and application of Article 5.3

Our analysis revealed that Aotearoa NZ’s application of Article 5.3 is largely confined to a single recommendation, which states that Parties should “ensure that…interactions” with tobacco industry actors “are conducted transparently” [35, Rec. 2.2]. Two government agencies, the Ministry of Health and New Zealand Customs Service (Customs), maintain public registers of meetings including with tobacco industry representatives [21,36]. Tobacco industry engagement with other government agencies appears to be unreported.

Our analysis of existing government responses to official information requests revealed that tobacco companies engaged with multiple government agencies and political entities other than Ministry of Health and Customs, thereby avoiding the only agencies applying Article 5.3. For instance, between 2018-2019, PMI directly approached government-funded hospitals offering its heated tobacco products free for cessation trials [37], and simultaneously lobbied a government advisory group established under the auspices of Treasury and Inland Revenue for preferential tax treatment of these products [38,39]. Existing documents revealed that a hospital doctor approached by PMI redirected them to the Ministry of Health, citing Aotearoa NZ’s obligations to Article 5.3 [37]. The correspondence also shows a Ministry of Health official declining a meeting with JUUL representatives, and recommending that the company “monitor the (Ministry of Health) website” for public consultation opportunities [37]. These are isolated examples of Recommendation 2.1 implementation, which advises Parties to restrict interactions with the tobacco industry to those “strictly necessary to enable them to effectively regulate the tobacco industry and tobacco products” [1 Recommendation 2.1].

Outside the Ministry of Health and Customs registers, overall awareness and application of Article 5.3 is inconsistent. In 2018, two government-funded hospitals invited the director of a research institute funded by Foundation for a Smoke-Free World (a research funding entity established and funded entirely by PMI) to speak at staff cessation training events [40–42]. When questioned by public health experts about this, one hospital chief executive stated, “the [hospital] does not have a policy on relations with the tobacco industry” [40]. Following subsequent media exposure of this, the Director-General of Health wrote to hospital chief executives to remind them of Article 5.3 obligations [43].

### Limited awareness among political actors

Political informants described limited awareness of Article 5.3, with one politician stating they were “happy to admit” their “ignorance”. WHO guidelines mandate awareness-raising across executive, legislative, and judicial branches regarding tobacco industry interference [1 Recommendation 1.1].

One politician involved in tobacco control policymaking stated they did not “recall being specifically lobbied by the tobacco industry” during tobacco-related policy events. The same informant then described email correspondence with “some within the tobacco industry” about “huge quantities of illegal tobacco” being imported. The industry actors provided the politician with “suggestions” for parliamentary questions to pose to government. Tellingly, the politician informant characterised this as being “spoken to” rather than “lobbied”:

> I’d have to go and look to that Convention that you’re referencing, but there probably is a distinction between being lobbied to say, “Hey, we want [people] to smoke” versus “Actually, there is some illegal behaviour happening at the border.” I’d probably make a quick distinction there. So, I have been lobbied, well not lobbied, I’ve been spoken to on that front. (Politician)

Article 5.3 does not make this distinction. The informant acknowledged the strategic nature of these communications, saying “I’m not dumb enough to ignore the fact that the tobacco industry doesn’t like competition [laughs].” Our analysis of policy submissions, parliamentary debates and ministerial questions revealed that illicit tobacco concerns have been repeatedly raised by industry and centre-right politicians against tobacco control measures; these arguments appear in discussions of point-of-sale display controls [44,45], plain packaging [46], excise tax increases [47,48], and vape controls [49]. Most recently, Prime Minister Christopher Luxon justified the 2024 policy repeal by claiming the measures would have created “an increased black market” [11].

Another politician involved in tobacco control policy spoke of the tobacco industry’s “unpopular” reputation but repeatedly argued that this should not exclude them from policy discussions, while also clarifying they had not personally met with the industry:

> I was always open to meet with [pause] I didn’t meet any tobacco companies - but I think, in a democracy you’ve got to at least engage with all sides of an argument. Even if it’s an unpopular side to a debate. (Politician)

When asked what guidance this informant received about interacting with industry interests, they stated:

> People give guidance around, there might be sometimes sensitivities, political sensitivities. For example, meeting with tobacco companies. Yeah, I think it’d be quite widely known that it’s not a good look… What would it look like [on the front page of the newspaper], the optics of it? I find that quite hard because my true nature is to listen to all sides of an argument… In our society, in our democracy, I don’t think it’s healthy to not sit down … I just don’t think that’s a healthy democracy. (Politician)

The politician alluded to the poor reputation of the tobacco industry and media optics rather than Aotearoa NZ’s obligations under Article 5.3.

### Limited oversight of tobacco industry influence

The narrowness of Article 5.3 implementation is a particular problem in the context of Aotearoa NZ’s existing transparency mechanisms. Our documentary analysis indicates the current systems provide inadequate oversight of tobacco industry influence, creating exploitable gaps that Article 5.3 is designed to address. The Official Information Act 1982 (OIA) [50] as well as the proactive disclosure of ministerial diary summaries are regularly used as exemplars of the nation’s open and transparent political system, and cited as the reason lobbying laws are unnecessary [51,52]. Yet a comparison against OECD lobbying transparency standards concluded its ability to provide insight into lobbying activities was limited [53]. For example, the Act does not extend to elected MPs, who are instead subject to a parliamentary information protocol allowing them to veto disclosure requests [54]. This suggests MPs operate outside both Article 5.3 protections and transparency mechanisms available for scrutinising tobacco industry-official interactions. This is a significant gap: MPs are key lobbying targets due to their roles in proposing and amending legislation, select committee processes, scrutinising government performance, and as the “pool of talent” from which Ministers are chosen [55]. A further limitation is that official information requests can be refused if information “cannot be made available without substantial collation or research” [50], a provision likely to impede comprehensive lobbying-related requests [53]. Ministerial diary summaries are similarly constrained. The published summaries record only formally scheduled ministerial meetings, excluding informal interactions, and Cabinet guidance contains no reference as to whether or how disclosure is monitored [56]. One journalist informant described the system as “better than nothing… but it’s voluntary, isn’t it?” and argued that “you’re only ever going to put something in the ministerial diary now that you’re comfortable with being made public.”

## DISCUSSION

This study demonstrates how tobacco industry interference operates in Aotearoa NZ, enabled by systematised limitations in Article 5.3 implementation. The institutional ethnographic approach provides insight into how policy and implementation gaps and the relative strength of governing texts influence lobbying activities, revealing mechanisms typically invisible in jurisdictions without regulated lobbying controls or disclosure. The patterns identified here suggest structural conditions under which policy reversals may be more feasible.

Aotearoa NZ’s approach primarily restricts protections to two agency-level meeting registers, a significant departure from comprehensive WHO guidelines. Studies from Uganda [57] and India, Bangladesh and Ethiopia [58] have documented similar patterns where Article 5.3 is viewed as solely a health ministry responsibility. This fragmented approach is exacerbated by Aotearoa NZ’s absence of lobbying laws, leaving it particularly exposed compared to many other Parties to the WHO FCTC who rely on existing mechanisms such as lobbying controls [4]. This narrow implementation creates exploitable vulnerabilities [4]. By confining Article 5.3 to the Ministry of Health and Customs, and by leaving other government bodies unprotected, Aotearoa NZ’s approach enables industry to avoid even this limited safeguard. Existing research highlights how the tobacco industry strategically targets non-health agencies to circumvent health ministry protections [4,59]. PMI’s 2014 corporate affairs presentation explicitly identifies moving “tobacco issues away from [Ministries of Health]” as central to “playing the political game” [60]. Our documentary analysis reveals PMI’s multifaceted influence strategy in Aotearoa NZ, with the company gaining access to local researchers and health providers through the its funding body, Foundation for a Smoke-Free World, while simultaneously lobbying government advisors for beneficial tax policies.

These structural vulnerabilities are compounded by an apparent lack of awareness of Article 5.3 obligations among elected officials, including those directly involved in tobacco control decision-making, a pattern also seen among members of the European Parliament [5]. Our findings indicate that decisions on tobacco industry engagement are coordinated through considerations of media optics and reputational concerns, and what constitutes industry engagement remains undefined. The politician who framed being “spoken to” about the industry’s illicit tobacco trade concerns as distinct from being “lobbied” to promote products exemplifies how these interactions are normalised as procedurally legitimate despite Article 5.3 protections.

The tobacco industry routinely and deliberately expresses concerns about illicit trade as a means to counter tobacco control regulations, while sponsoring research supporting this position [61,62]. In Aotearoa NZ, this argument has been deployed repeatedly against strengthened controls, with effectiveness evident in its recent use by the Prime Minister in support for the policy repeals [11]. Without comprehensive understanding of Article 5.3 or industry tactics, political actors can become conduits for industry arguments while believing they are addressing legitimate policy concerns. This lack of understanding, manifested in one politician’s depiction of interaction restrictions as barriers to hearing “all sides of an argument,” reflects how Article 5.3 can be misconstrued as anti-democratic. This aligns with strategies whereby tobacco companies employed “good governance” rhetoric to reframe Article 5.3 protections as being at odds with fair and open consultative policymaking [63].

Hawkins and Holden [5] observed general reluctance in the European Parliament to restrict or disclose Members’ interactions with the tobacco industry given their roles as legislators and constituent representatives. Australia is one of few countries applying recommendations which reflect WHO guidelines: Australian Department of Health guidelines establish that ‘public officials’ subject to Article 5.3 obligations include MPs and their staff, government employees, contractors and consultants across all government branches [64]. This suggests that legislative branch exclusion from Article 5.3 implementation represents a systematic global pattern rather than jurisdiction-specific challenges.

The limited visibility of political decision-making processes in Aotearoa NZ creates further weaknesses for observing industry influence: the OIA cannot adequately capture lobbying activity [53]; despite their crucial legislative responsibilities, the parliamentary information protocol allows MPs to veto information disclosure requests [54], creating another gap; and ministerial diary summaries provide limited and inconsistent records. Aotearoa NZ’s existing system gives the impression of openness without providing genuine transparency or accountability.

This lack of scrutiny is consequential. Internal company documents show that tobacco and vaping companies directly target MPs in Aotearoa NZ to prevent strengthened tobacco control [65–67]. Internal emails from JUUL reveal the company strategically identified and targeted specific Ministers, MPs and officials seen as “potential allies” in 2019 as Parliament moved to introduce vaping regulations [68,69]. While officials declined to engage, JUUL successfully connected with MPs [68]. JUUL’s direct access to MPs only ended in July 2019 when the Ministry of Health issued a directive requiring that all further communication with JUUL go through them [70], suggesting that MPs were unaware of Article 5.3 protections. This reflects a broader reaction pattern of in Aotearoa NZ; Article 5.3 protections have been applied after instances of industry contact came to light, rather than institutionally embedded, as recommended within the WHO FCTC.

Our findings highlight the need for reform. Aotearoa NZ and other Parties should consider comprehensive Article 5.3 implementation codification across all government branches. The Tobacco Transparency Bill, a Private Members’ Bill introduced in May 2025 in response to the 2024 policy reversals, proposes to address several of these deficits in Aotearoa NZ by codifying key aspects of Article 5.3 in law [71]. However, several gaps remain. For example, the Bill excludes the legislative branch (MPs), leaving a crucial industry influence pathway intact. Establishing clear protocols for restricting interactions that include MPs would enable all political actors to understand and comply with Article 5.3 obligations [1]. In light of the disproportionate impacts of tobacco on Māori and Pacific peoples, explicit incorporation of Te Tiriti o Waitangi commitments, equity specific targets, and Māori and Pacific leadership in all tobacco control legislation would help to ensure equity is not treated as optional.

### Limitations

Aotearoa NZ’s absence of lobbying disclosure requirements prevented independent verification of the full scope of tobacco industry activities, with findings necessarily reliant on informant accounts and publicly available documents. Voluntary participation may have created selection bias toward those more sympathetic to lobbying protections, limiting the generalisability of findings. The absence of Ministry of Health tobacco control officials participating is a key limitation, requiring greater reliance on the experiences of public health advocates and documentary analysis.

## CONCLUSION

Despite its international standing as a tobacco control leader, this research shows that Aotearoa NZ’s implementation of Article 5.3 of the WHO FCTC is limited and reactive. Compounded by a lack of lobbying laws and ill-equipped transparency mechanisms, these policy and implementation failures have produced institutional and political vulnerabilities that may enable tobacco industry interference in health policy. Comprehensively embedding whole-of-government protections that uphold Article 5.3 by Aotearoa NZ and other Parties is urgent and necessary to safeguard policy decisions from pervasive tobacco industry influence.

## ACKNOWLEDGEMENTS

We gratefully acknowledge the late Professor Kypros Kypri for his substantial contributions to the study’s conception, design, analysis and supervision. We also gratefully acknowledge the study participants who shared their insights experiences and time for the purposes of this research. Any errors in interpretation are the responsibility of the authors.

## AUTHOR CONTRIBUTIONS

M-J.G.: Conceptualization, Data curation, Formal analysis, Investigation, Methodology, Project administration, Writing – original draft and review & editing. J.W.: Conceptualization, Methodology, Supervision, Writing – review & editing. P.A.: Conceptualization, Funding acquisition, Methodology, Supervision, Writing – review & editing. V.N.: Conceptualization, Supervision, Writing – review & editing. E.W.: Conceptualisation, Supervision, Writing – review & editing.

## ETHICS

Ethical approval was granted by the University of Auckland Human Participants Ethics Committee (UAHPEC21554) from November 2020 until November 2023.

## DATA AVAILABILITY

The data underlying this article cannot be shared for the privacy of the individuals that participated in this study.

